# A household case evidences shorter shedding of SARS-CoV-2 in naturally infected cats compared to their human owners

**DOI:** 10.1101/2020.10.31.20220608

**Authors:** Víctor Neira, Bárbara Brito, Belén Agüero, Felipe Berrios, Valentina Valdés, Alberto Gutierrez, Naomi Ariyama, Patricio Espinoza, Patricio Retamal, Edward C. Holmes, Ana S. Gonzalez-Reiche, Zenab Khan, Adriana van de Guchte, Jayeeta Dutta, Lisa Miorin, Thomas Kehrer, Nicolás Galarce, Leonardo I. Almonacid, Jorge Levican, Harm van Bakel, Adolfo García-Sastre, Rafael A. Medina

## Abstract

Severe acute respiratory syndrome coronavirus 2 (SARS-CoV-2) has been detected in domestic and wild cats. However, little is known about natural viral infections of domestic cats, although their importance for modeling disease spread, informing strategies for managing positive human-animal relationships and disease prevention. Here, we describe the SARS-CoV-2 infection in a household of two human adults and sibling cats (one male and two females) using real-time RT-PCR, an ELISA test, viral sequencing, and virus isolation. On May 2020, the cat- owners tested positive for SARS-CoV-2. Two days later, the male cat showed mild respiratory symptoms and tested positive. Four days after the male cat, the two female cats became positive, asymptomatically. Also, one human and one cat showed antibodies against SARS-CoV-2. All cats excreted detectable SARS-CoV-2 RNA for a shorter duration than humans and viral sequences analysis confirmed human-to-cat transmission. We could not determine if cat-to-cat transmission also occurred.

**Article Summary Line:** SARS-CoV-2 in naturally infected cats present a shorter shedding pattern compared to their owners.

## Introduction

In December 2019, severe acute respiratory syndrome-related coronavirus 2 (SARS-CoV-2) emerged as a new human pathogen in Wuhan, Hubei province, China, likely due to a cross-species transmission event between humans and an unknown animal reservoir(1). Since then, SARS-CoV-2 has been efficiently transmitted between humans, causing a pandemic with devastating consequences for human health, societies and economies globally(2).

There is also now a growing body of data indicating that the virus is able to infect companion animals and other animal species(1,3). To date, naturally occurring cases officially reported to the World Organization for Animal Health (OIE) and to the United States Department of Agriculture include 14 cases in dogs in the US, Hong Kong, and Japan, and 25 cases in cats in the US, Hong Kong, France, Belgium, Germany, Russia, Spain and the UK(4). Additionally, cases in farmed mink from the Netherlands, Spain, US, and Denmark, as well as tigers and lions from a New York zoo in the US, have also been reported(4–6). Dogs that have been experimentally infected show low viral loads and do not observably transmit SARS-CoV-2 between dogs(1). Among domestic animals, natural infection and experimental data suggest that felids and mustelids are more susceptible to SARS-CoV-2 infection than canines(4,7,8). Companion animals are considered as protective factors against loneliness and as a source of social and emotional support for people during difficult situations or disease(9–11).

Understanding how and when SARS-CoV-2 is transmitted in domestic animals is critical for informing decisions for control and prevention of infections. Previous studies of SARS-CoV- 2 transmission in cats have been in laboratory settings. Cats experimentally infected with SARS-CoV-2 can efficiently replicate and transmit the virus to other cats housed in adjacent cages, suggesting respiratory transmission. In these species, the virus replicates mostly in the upper airways. Juvenile compared to sub-adult cats have more severe outcomes(7,12). However, nothing is known about patterns of viral transmission under natural conditions in households in which humans inhabits with companion cats. Key questions involve determining the dynamics of virus infection including the duration of viral excretion.

Beginning in May 2020, we conducted active surveillance in Santiago City, Chile, to determine the potential infection of companion animals in households in which the humans had tested positive to SARS-CoV-2. At that time, Santiago was experiencing daily reports of >3000 SARS-CoV-2 human cases. From a survey of 12 households, we found two containing companion animals that tested positive for SARS-CoV-2. One household had two cats, one of which presented antibodies to SARS-CoV-2. The other household was home to three 10-year-old sibling cats: all three were identified as having a SARS-CoV-2-positive infection. Herein, we describe a case-study of naturally occurring SARS-CoV-2 infection based on the three-cat household that also included two human adults aged in their 30s and 60s years old who were both positive to SARS-CoV-2. The data generated provide a better understanding of infection dynamics in cats, one of the most important SARS-CoV-2 animal natural hosts documented to date.

## Methods

### Case description and specimen collection

Between May and September 2020, we collected animal companion samples from 12 households with confirmed human SARS-CoV-2 cases in Santiago City, Chile. These samples were from 17 cats and 10 dogs (Supplementary information). Of the 12 households sampled, two presented animals positive for SARS-CoV-2. In one household, one of two cats had SARS-CoV- 2 antibodies measured by ELISA. In the other household, three out of three cats were positive for SARS-CoV-2 by rtRT-PCR. We followed the second-mentioned household for 40 days to examine SARS-CoV-2 infection dynamics. This case is described below.

On May 2020 a COVID-19 diagnosis was confirmed by SARS-CoV-2 rtRT-PCR for the humans in the studied household – a male of around 30 years old and a female of around 60 years old. The residents owned three 10-years-old sibling cats, 1 male (cat 1) and 2 females (cat 2 & 3). Of the three cats, cat 1 had the closest contact with the humans, sharing human 1’s bed. Humans and cats were recruited to this case study. From humans, nasal swabs, sputum, and fresh fecal samples were collected at intervals of 4 to 7 days. From cats, nasal swabs and fresh fecal samples were collected at intervals of 1 to 3 days until rtRT-PCR was negative for SARS-CoV-2. At day 0 (first sampling date) a nasal swab was collected from cat 1 and fresh feces were collected from all 3 cats. Cat 1 tested positive for SARS-CoV-2 by rtRT-PCR in both nasal (Ct=31) and fecal sample (Ct=33), and cat 2 and 3 tested negative. In cats, at the last sampling, serum samples were also collected. Nasal samples were collected using flocked swabs; fresh feces were stored in ziplock bags(13–15) and deposited in viral transport media.

The owners provided written consents at the time of the sample collection. For all the study, human sampling was performed by nurses and animal sampling was performed by veterinarians. Humans samples were tested at the Molecular Virology Laboratory, Pontificia Universidad Católica de Chile and animal samples were tested at the Animal Virology Laboratory, Universidad de Chile, La Pintana, Chile. The animal study was reviewed and approved by the Institutional Animal Care and Use Committee of Universidad de Chile, under protocol number 20370–VET–UCH, Biosafety Committee of Facultad de Ciencias Veterinarias y Pecuarias, Universidad de Chile under protocol number 161 and the human study was reviewed and approved under protocol 16-066 Scientific Ethical Committee on Health Sciences of Pontifical Universidad Católica de Chile.

### Real time RT-PCR

RNA from the samples was extracted using TRIzol™ Reagent (Invitrogen™, Carlsbad, CA, USA) following the instructions recommended by the manufacturer and eluted into 50 µL. The extracted RNA was subjected to rtRT-PCR to amplify a portion of ORF1b SARS-CoV- 2(16). This PCR was previously validated at both laboratories independently, and is one of the RT-PCRs recommended by the WHO(17) to diagnose COVID-19. Positive and negative controls for PCR and extraction were included in each run.

### Serological testing

Serum samples were tested via a commercial ELISA test for SARS-CoV-2 antibody detection, the ID SCREEN® SARS-COV-2 Double Antigen Multi Species(18) (IDVET, Grabels, FRANCE).

### Viral isolation

Viral isolation was attempted in Vero E6 cells at Icahn School of Medicine at Mount Sinai, New York. Cells were cultured in minimum essential medium eagle 10% fetal bovine serum, 100 IU/mL penicillin, and 100 µg/mL streptomycin. Cells were seeded in 12-well culture plates and cultured at 37°C with 5% CO_2_ for 24-48h. Cells were rinsed and inoculated with 300 µl of the direct sample and 1:10 and 1:100 dilutions. Mock-inoculated cells were used as negative controls. Cells were incubated at 37°C with 5% CO_2_ and monitored daily for cytopathic effect (CPE) for five days. Cell cultures with no CPE were frozen, thawed, and subjected to three blind passages with inoculation of fresh Vero E6 cell cultures with the lysates as described above.

### Viral sequencing

A selection of 25 rtRT-PCR positive specimens from humans and cats was submitted to the Icahn Institute for Data Science and Genomic Technology, Icahn School of Medicine at Mount Sinai, New York, NY for whole genome sequencing. Selection criteria were based on the cycle threshold values and inclusion of all cats and humans. The whole genome was amplified with a modified version of the Artic Consortium protocol(19) with custom tiling primers as previously described(20). After amplification, viral sequencing was attempted on 12 positive samples, four human and eight from cat. Purified amplicons were used for library preparation with Nextera XT DNA Sample preparation Kit (Illumina, FC-131-1096) and sequenced on an Illumina MiSeq instrument (2×150 nt). SARS-CoV-2 genomes were assembled and annotated using a reference-based analysis pipeline as previously described(20).

### Sequence data analysis

Six SARS-CoV-2 genome sequences were analyzed. These sequences were obtained from the clinical nasopharyngeal specimens of two humans that reside in the household: Human 1 (hCoV-19/Chile/Santiago/op31d5/2020; Human 1) and Human 2 (hCoV- 19/Chile/Santiago/op32d10/2020; (Human 2), and four sequences from their cats - two of these were collected from nasal swabs of cat 2, and a nasal sample from cat 3. Completeness of sequencing coverage and mutations were estimated by comparing the alignment with the reference sequence Wuhan-Hu-1 (GenBank accession NC_045512.2). Sequences were aligned to a set of reference SARS-CoV-2 sequences including all Chilean sequences submitted to GISAID(20) (n=167), sequences with highest similarity deposited on GenBank identified using BLAST(21) (n=337), and a total of 1000 sequences randomly selected from the GISAID repository using FastaUtils package in R(22). The sequences were aligned using MAFFT v7.402 using CIPRESS computational resources(23). The aligned sequences were visualized in AliView v1.26(24). Duplicate sequences were removed and non-coding regions, 5’ and 3’ UTR were trimmed. A phylogeny was estimated using the maximum likelihood method available in IQ-TREE v1.6.7 employing the GTR+F+I+Г_4_ model of nucleotide substitution with 1000 bootstrap replications. The phylogenetic tree was visualized using Figtree. SARS-CoV-2 lineages were identified using the Pangolin COVID-19 Lineage Assigner web application(25).

## Results

### Naturally infected cats excreted SARS-CoV-2 RNA in a different pattern to that of humans

To determine the duration and dynamics of SARS-Cov-2 RNA detection in cat excretions under natural conditions, we assessed, by real time RT-PCR (rtRT-PCR), the presence of SARS-CoV-2 RNA in nasal and fecal samples from three companion cats over a 40-day time period.

The cats acquired the SARS-CoV-2 infection naturally (rather than experimentally) and were residing in their usual household during the sampling period. Cat 2 had a pre-existing chronic rhinosinusitis as comorbidity. The household included two humans, and both were confirmed positive for SARS-CoV-2 before sampling of the cats commenced. Nasal, sputum, and fecal samples were taken from the humans and also assessed by rtRT-PCR for the presence of SARS-CoV-2 RNA over the time period in which the cats were sampled. All three cats had detectable viral RNA in both nasal swabs and fecal samples, with cycle threshold (Ct) values over 21.9 (Figure 1). In both humans, nasal and sputum samples showed Ct values ranging from 26 to 40. However, all human fecal samples showed negative rtRT-PCR results. The duration for which SARS-CoV-2 RNA could be detected in cat samples varied between the three cats sampled (5 days for cat 1, 8 days for cat 3, 17 for cat 2). Even at the extreme end of the range, the duration of detection was considerably shorter than the human samples. SARS-CoV-2 RNA was detected in the human samples up to 25 days after the first RNA detection. The dynamics of detectable viral RNA in the cat samples were more erratic compared to that for detectable viral RNA in humans from the same household. For example, a female cat (cat 2) presented an intermittent RNA detection for at least 17 days (Supplementary information).

**Figure 1.**
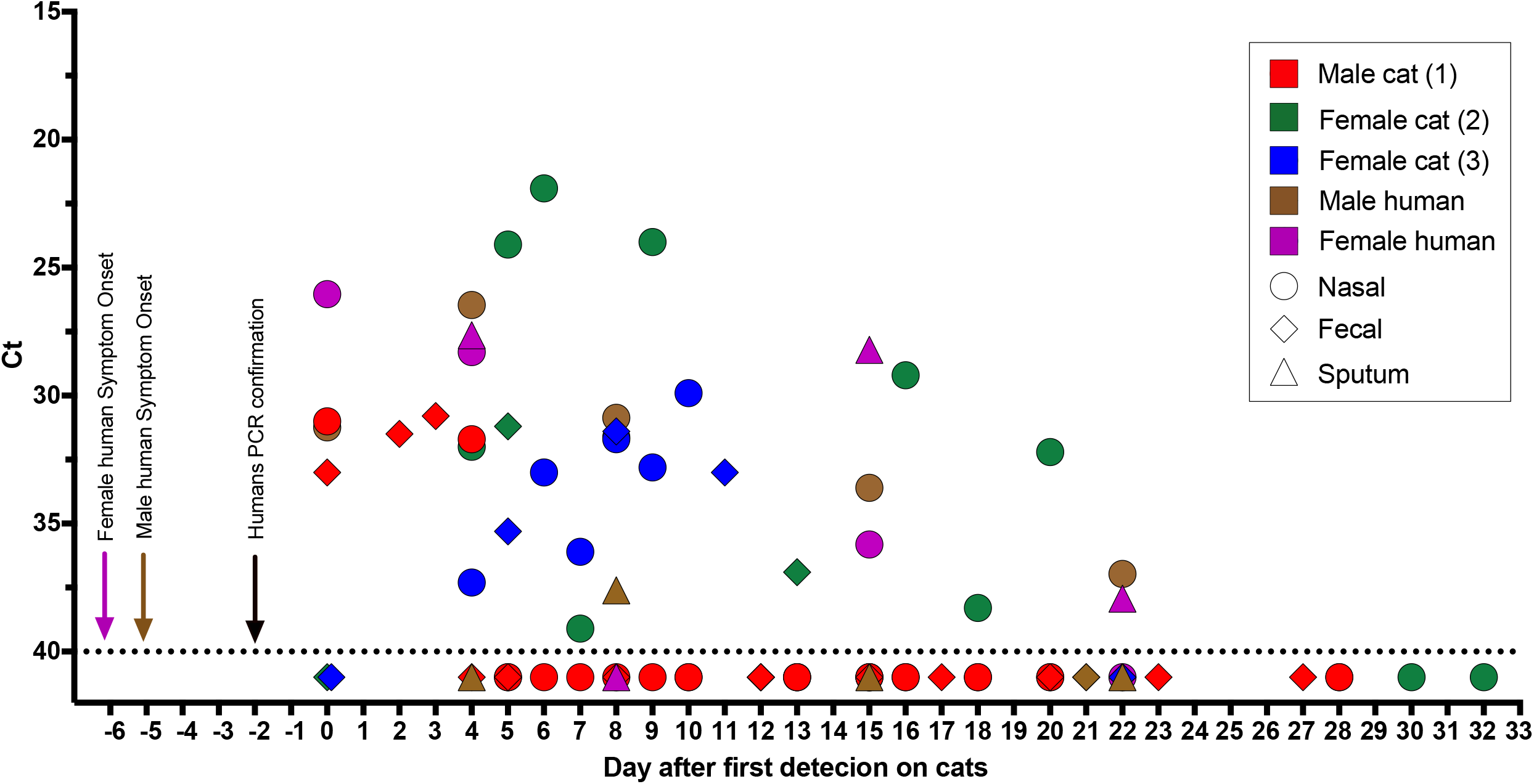
RNA detection of SARS-CoV-2. The horizontal axis represents the sampling date. Nasal swabs (circle), fecal samples (diamond), and sputum (triangle) samples are colored per individual. Negative samples are illustrated as Ct 41 (below the dotted line). Arrows with the corresponding caption colors indicate the day of symptom onset and the date of the SARS-CoV- 2 diagnosis of humans.

To assess the infection of the cats and humans, we made clinical observations for symptoms of SARS-CoV-2 infection and performed serological testing. Cat 1 presented with mild clinical signs including dullness, lethargy, and coughing without fever. Cats 2 and 3 were asymptomatic. On the last sampling on day 33 after first detection on cats, cat 2 tested positive for SARS-CoV-2 antibodies by ELISA. Both humans displayed COVID-19 symptoms including fever, lethargy, and coughing. However, the female human was more clinically affected and was the only human that tested positive for SARS-CoV-2 antibodies by ELISA starting at day 4 after detection on cats.

Multiple attempts to isolate the virus from low Ct (high levels of SARS-CoV-2 RNA; 21.9 −24.1) samples were made without success.

### Human and cat origin SARS-CoV-2 genome sequences are identical

Of the specimens sent for genome sequencing one had coverage of 69·3% (human 2, day 10, nasopharyngeal) and five had a genome coverage >98% (three nasal swabs from cat 2, one nasal swab from cat 3, and one nasopharyngeal from human 1) (Appendix Figure). The remaining sequenced samples had a coverage <50% and were not included in the analyses. Sequences were deposited in GenBank (Accession numbers MW064259- MW064264). Using the Pangolin tool(25) all sequences were assigned to lineage B.1.1, a major European lineage that was exported to the rest of the world from Europe. Of the 178 Chilean sequences included in this analysis (from GISAID and GenBank), 34 belong to this lineage.

We conducted a phylogenetic analysis to investigate any relationship between the SARS-CoV-2 RNA excreted by different cats and humans within the household. All six sequences obtained fell into a monophyletic group defined by a C25344T (S1261F/Spike) nucleotide substitution (Figure 2). Notably, there were no nucleotide differences between the viral sequences obtained from cats and one of the household humans (hCoV- 19/Chile/Santiago/op31d5/2020; Human). The viral sequence from the female human (hCoV- 19/Chile/Santiago/op32d10/2020; (Human 2) contained two polymorphisms compared to the rest of the sequences (C11511T/T3749I/nsp6) and (C13740T/syn/RdRp) but fell in the same group. An epidemiologically unrelated Chilean SARS-CoV-2 sequence (GISAID accession number EPI_ISL_468748) collected in the same city on April was also grouped within this cluster of 6 sequences.

**Figure 2.**
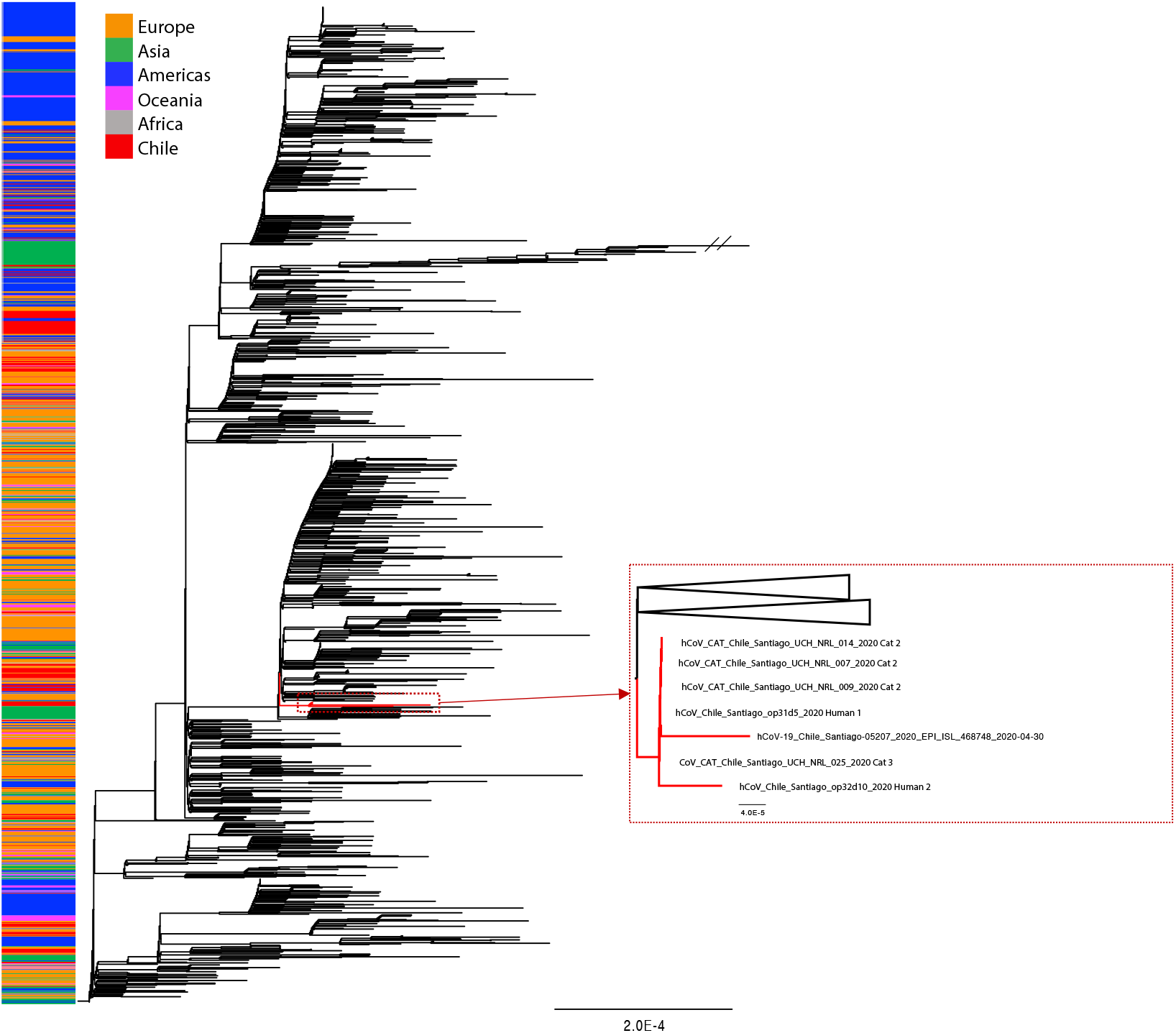
Phylogenetic tree estimated with the all available Chilean SARS-CoV-2 sequences and a random sample of available background sequences. Near to complete genome sequences of cat 2 (n=3), cat 3 (n=1), and both owners formed a monophyletic group. An additional human sequence collected in the same time period and area similarly fell into this cluster. The phylogeny was estimated using the maximum likelihood method available in IQ-TREE v1.6.7 employing the GTR+F+I+Г4 model of nucleotide substitution with 1000 bootstrap replications.

## Discussion

Several reports have confirmed the susceptibility of cats to SARS-CoV-2 infection in natural conditions and viral transmissibility between cats under experimental conditions(12,26,27). However, these do not provide insight into the infection dynamics of cats in natural settings. This is the first report to describe the RNA excretion profiles of SARS-CoV-2 in naturally infected cats. The viral RNA in samples may indirectly denote SARS-CoV-2 viral excretion and can serve, with limitations, as a proxy for active viral replication. Indeed, SARS-CoV-2 antibodies detected by ELISA confirmed the infection in cat 2. Our household study thus provides preliminary guidance for managing the care and quarantine of cats in households in which SARS-CoV-2 is present.

The results, particularly the similarity of genome sequences depicted in the phylogenetic analysis, strongly support the idea that SARS-CoV-2 can be transmitted between humans and cats living in the same household. It is suspected that the humans were infected following exposure to SARS-CoV-2-positive neighbors days before. We think it likely that cat 1 was the first infected of the three cats because this animal had closer contact than the other two cats with human 1 (male), sharing his bed. This is supported by our observation that viral RNA from human 1 and the cats had identical sequences whereas the female human’s viral RNA differed by two polymorphisms. Transmission between humans and cats in the same household accords with the literature on viral infections in animals. Several animal species are permissive for virus infection and replication in the upper respiratory tract(1) and it has been suggested that the species barrier of SARS-CoV-2 might be weak since the ACE2 host receptor may allow virus attachment even after some amino acid changes(28). SARS-CoV-2 can be transmitted to animals by direct or indirect contact with contaminated surfaces or excretions(29). For cats(1) and ferrets(30) respiratory transmission can be mediated by droplets or aerosols. Our findings of positive SARS-CoV-2 RNA detection in nasal samples (ranging from 5-17 days) and in fecal samples (ranging from 4-10 days) in all three cats, further support recommendations for avoiding close contact with companion animals when an infection is suspected in owners or family members.

Whether all cats in the study household were infected from humans, or whether some transmission occurred between animals, cannot be determined. Nevertheless, the possibility of some transmission between cats in the household is suggested by the 4-day delay in cats 2 and 3 testing positive to SARS-CoV-2 by rtRT-PCR compared to cat 1. All three cats had direct contact with each other. Transmission between cats in the household would be consistent with the findings of Shi et al. (2020) who studied transmission between cats in an experimental setting^7^. In that study, inoculated and infected sub-adult cats were placed next to susceptible cats in an adjacent cage (non-direct contact). Of the three replicates, one exposed cat gave rise to a positive rtRT-PCR on the 5th day post-exposure(7). To date, however, we have only identified three rtRT-PCR-positive cats, with one of these also positive by ELISA, and one cat of another household similarly positive by ELISA. Consequently, it is likely that cat-to-cat transmission events are not as common as human-to-human transmission.

In summary, at present, there is insufficient data to identify and prioritize the drivers for the infection of companion animals(31). Reported cases in animals correspond to independent and sporadic events. The findings presented here show that cats seemingly have different and shorter patterns of excretion for SARS-CoV-2 RNA compared to humans. These results highlight a need for large-scale epidemiological analysis of SARS-CoV-2 infection dynamics to support the establishment of preventive measures in real-life human-animal relationships.

## Data Availability

Sequences that support the results of this study have been deposited at GenBank with accession numbers MW064259 - MW064264.

## Acknowledgments

This study was partly funded by the Programa Fondecyt de Iniciación N° 11170877 and Proyecto FIV to VN; the Programa Beca Magister Nacional de ANID N°22190312 to NA. We thank Randy Albrecht for support with the BSL3 facility and procedures at the ISMMS, and Richard Cadagan for technical assistance. These studies were partly supported by CRIP (Center for Research for Influenza Pathogenesis), a NIAID supported Center of Excellence for Influenza Research and Surveillance (CEIRS, contract # HHSN272201400008C) to VN, RAM, HVB, and AG-S; by the generous support of the JPB Foundation, the Open Philanthropy Project (research grant 2020-215611 (5384)); and by anonymous donors to AG-S. This research was also partly supported by the Office of Research Infrastructure of the National Institutes of Health (NIH) under award numbers S10OD018522 and S10OD026880. ASG-R. was further supported by a Robin Chemers Neustein Postdoctoral Fellowship Award. Human protocols and the study set-up used for this study were based on influenza virus studies established in part with the support of the FONDECYT 1161971 grant to RAM and by grant ACT 1408 to RAM and VN from the Agencia Nacional de Investigación y Desarrollo (ANID) from Chile. We gratefully acknowledge the GISAID EpiFlu™ Database, laboratories, and original source of data of Chilean SARS-CoV- 2 sequences (supplementary information), especially to the Facultad de Medicina Pontifical Universidad Católica de Chile and the Center for Mathematical Modeling and the Center for Genome Regulation at Universidad de Chile, source of the EPI ISL 468748.

## Disclaimers

The authors declare no competing interest.

## Author Bio

Dr. Neira is assistant professor at the Animal Virology Unit, Facultad de Ciencias Veterinarias y Pecuarias, Universidad de Chile in Santiago. His research interests include understanding the diversity, transmission, and ecology of emerging and reemerging viruses in wildlife, companion, and livestock animals.

**Appendix Figure**. Representation of the genome coverage and mutations with respect to Wuhan-Hu-1 reference sequence NC_045512.2. Dark grey shaded areas indicate no coverage. Blue and red lines indicate synonymous and non-synonymous substitutions respectively. The specific nucleotide substitutions and amino acid changes are indicated at the top and bottom of the figure.

**Supplementary information 1**. Table 1. Summary of cases on study.

**Supplementary information 2**. Table 2. Individual results.

**Supplementary information 3**. Acknowledge to GISAID

## Notes

### Competing Interest Statement

The authors have declared no competing interest.

### Funding Statement

This study was partly funded by the Programa Fondecyt de Iniciacion N 11170877 and Proyecto FIV to VN; the Programa Beca Magister Nacional de ANID N 22190312 to NA. These studies were partly supported by CRIP (Center for Research for Influenza Pathogenesis), a NIAID supported Center of Excellence for Influenza Research and Surveillance (CEIRS, contract # HHSN272201400008C) to VN, RAM, HVB, and AG-S; by the generous support of the JPB Foundation, the Open Philanthropy Project (research grant 2020 215611 (5384)); and by anonymous donors to AG-S. This research was also partly supported by the Office of Research Infrastructure of the National Institutes of Health (NIH) under award numbers S10OD018522 and S10OD026880. ASG-R. was further supported by a Robin Chemers Neustein Postdoctoral Fellowship Award. Human protocols and the study set-up used for this study were based on influenza virus studies established in part with the support of the FONDECYT 1161971 grant to RAM and by grant ACT 1408 to RAM and VN from the Agencia Nacional de Investigacion y Desarrollo (ANID) from Chile.

### Author Declarations

The animal study was reviewed and approved by the Institutional Animal Care and Use Committee of Universidad de Chile, under protocol number 20370 VET UCH, Biosafety Committee of Facultad de Ciencias Veterinarias y Pecuarias, Universidad de Chile under protocol number 161 and the human study was reviewed and approved under protocol 16066 Scientific Ethical Committee on Health Sciences of Pontifical Universidad Catolica de Chile.

## References

1. Shi J, Wen Z, Zhong G, Yang H, Wang C, Huang B, et al. Susceptibility of ferrets, cats, dogs, and other domesticated animals to SARS–coronavirus 2. Science (80-). 2020 Apr;eabb7015.

2. Liu T, Liang W, Zhong H, He J, Chen Z, He G, et al. Risk factors associated with COVID-19 infection: a retrospective cohort study based on contacts tracing. Emerg Microbes Infect. 2020 Jan;9(1):1–31.

3. Sit THC, Brackman CJ, Ip SM, Tam KWS, Law PYT, To EMW, et al. Infection of dogs with SARS-CoV-2. Nature. 2020 May;1–6.

4. 55 USDA APHIS. USDA APHIS | Confirmed cases of SARS-CoV-2 in Animals in the United States. 2020.

5. McAloose D, Laverack M, Wang L, Killian ML, Caserta LC, Mitchell PK, et al. From people to Panthera: Natural SARS-CoV-2 infection in tigers and lions at the Bronx Zoo. bioRxiv. 2020 Jul;2020.07.22.213959.

6. 57 Events in animals: OIE - World Organisation for Animal Health.

7. Shi J, Wen Z, Zhong G, Yang H, Wang C, Huang B, et al. Susceptibility of ferrets, cats, dogs, and other domesticated animals to SARS-coronavirus 2. Science. 2020 May;368(6494):1016–20.

8. Sit THC, Brackman CJ, Ip SM, Tam KWS, Law PYT, To EMW, et al. Infection of dogs with SARS-CoV-2. Nature. 2020 May;1–3.

9. Wood L, Martin K, Christian H, Nathan A, Lauritsen C, Houghton S, et al. The Pet Factor - Companion Animals as a Conduit for Getting to Know People, Friendship Formation and Social Support. Uchino BN, editor. PLoS One. 2015 Apr;10(4):e0122085.

10. Oliva JL, Johnston KL. Puppy love in the time of Corona: Dog ownership protects against loneliness for those living alone during the COVID-19 lockdown. Int J Soc Psychiatry. 0 2020 Jul;002076402094419.

11. Bowen J, García E, Darder P, Argüelles J, Fatjó J. The effects of the Spanish COVID-19 lockdown on people, their pets and the human-animal bond. J Vet Behav. 2020 Jun;

12. Halfmann PJ, Hatta M, Chiba S, Maemura T, Fan S, Takeda M, et al. Transmission of SARS-CoV-2 in Domestic Cats. N Engl J Med. 2020 May;

13. Neira V, Allerson M, Corzo C, Culhane M, Rendahl A, Torremorell M. Detection of influenza A virus in aerosols of vaccinated and non-vaccinated pigs in a warm environment. Huber VC, editor. PLoS One. 2018 May;13(5):e0197600.

14. Agüero B, Mena J, Berrios F, Tapia R, Salinas C, Dutta J, et al. First report of Porcine Respirovirus 1 in South America. Vet Microbiol. 2020 May;108726.

15. Tapia R, García V, Mena J, Bucarey S, Medina RA, Neira V. Infection of novel reassortant H1N2 and H3N2 swine influenza A viruses in the guinea pig model. Vet Res. 2018 Dec;49(1):73.

16. Chu DKW, Pan Y, Cheng SMS, Hui KPY, Krishnan P, Liu Y, et al. Molecular Diagnosis of a Novel Coronavirus (2019-nCoV) Causing an Outbreak of Pneumonia. Clin Chem. 2020;66(4):549–555.

17. 68 OMS. Molecular assays to diagnose COVID-19: Summary table of available protocols. 2020.

18. 69 IDVET. ID Screen® SARS-CoV-2 Double Antigen Multi-species - IDVet. 2020.

19. Sevinsky J. SARS-CoV-2 Sequencing on Illumina MiSeq Using ARTIC Protocol: Part 1 - 0 Tiling PCR. 2020 Apr;

20. Gonzalez-Reiche AS, Hernandez MM, Sullivan MJ, Ciferri B, Alshammary H, Obla A, et al. Introductions and early spread of SARS-CoV-2 in the New York City area. Science (80-). 2020 Jul;369(6501):297–301.

21. Altschul SF, Gish W, Miller W, Myers EW, Lipman DJ. Basic local alignment search tool. J Mol Biol. 1990 Oct;215(3):403–10.

22. 73 R Core Team. R: A language and environment for statistical computing. Vienna, Austria: R Foundation for Statistical Computing,; 2013.

23. Miller MA, Pfeiffer W, Schwartz T. The CIPRES science gateway. In: Proceedings of the 1st Conference of the Extreme Science and Engineering Discovery Environment on Bridging from the eXtreme to the campus and beyond - XSEDE ‘12. New York, New York, USA: ACM Press; 2012. p. 1.

24. Larsson A. AliView: a fast and lightweight alignment viewer and editor for large datasets. Bioinforma Appl. 2014;30(22):3276–8.

25. Rambaut A, Holmes EC, Hill V, OToole A, McCrone J, Ruis C, et al. A dynamic nomenclature proposal for SARS-CoV-2 to assist genomic epidemiology. bioRxiv. 2020 Apr;2020.04.17.046086.

26. Deng J, Jin Y, Liu Y, Sun J, Hao L, Bai J, et al. Serological survey of SARS_JCoV_J2 for experimental, domestic, companion and wild animals excludes intermediate hosts of 35 different species of animals. Transbound Emerg Dis. 2020 May;tbed.13577.

27. Zhang Q, Zhang H, Huang K, Yang Y, Hui X, Gao J, et al. SARS-CoV-2 neutralizing 0 serum antibodies in cats: a serological investigation. bioRxiv. 2020 Apr;2020.04.01.021196.

28. Zhai X, Sun J, Yan Z, Zhang J, Zhao J, Zhao Z, et al. Comparison of SARS-CoV-2 spike protein binding to ACE2 receptors from human, pets, farm animals, and putative intermediate hosts. J Virol. 2020 May;94(15).

29. Sarkar J, Guha R. Infectivity, virulence, pathogenicity, host-pathogen interactions of SARS and SARS-CoV-2 in experimental animals: a systematic review. Veterinary Research Communications. Springer; 2020. p. 1.

30. Richard M, Kok A, de Meulder D, Bestebroer TM, Lamers MM, Okba NMA, et al. SARS-CoV-2 is transmitted via contact and via the air between ferrets. Nat Commun. 2020 Dec;11(1).

31. Saegerman C, Bianchini J, Renault V, Haddad N, Humblet M. First expert elicitation of knowledge on drivers of emergence of the COVID_J19 in pets. Transbound Emerg Dis. 2020 Jul;tbed.13724.

