## Supplementary figures and images for "A household case evidences shorter shedding of SARS-CoV-2 in naturally infected cats compared to their human owners"

### Supplementary Figure 1

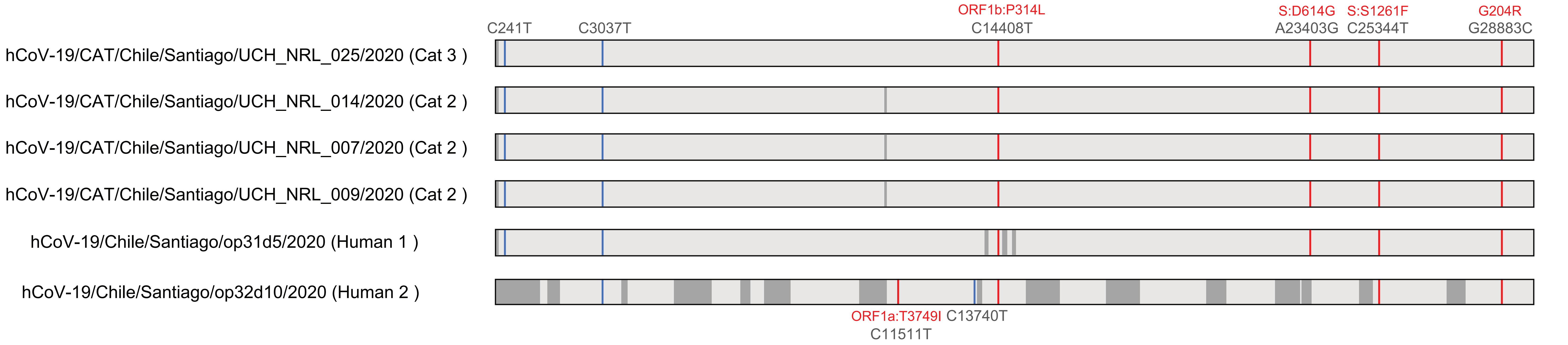
